# Evaluating Spiking and Non-Spiking Neural Networks for Colorectal Serrated Polyp Subtype Classification

**DOI:** 10.64898/2026.01.24.26344766

**Authors:** Nickolas Littlefield, Riyue Bao, Rong Xia, Qiangqiang Gu

## Abstract

Image classification on digital pathology images relies heavily on convolutional neural networks (CNNs), yet the behavior of alternative neural computing paragigms in this domain remains insufficiently characterized. Spiking neural networks (SNNs), which process information through event-driven spike-based dynamics, have recently become trainable at scale but have not been evaluated under standardized colorectal pathology benchmarks. This study presents the first controlled comparison of SNNs and CNNs on the Minimalist Histopathology Image Analysis (MHIST) Dataset, a widely used publicly available benchmark designed for reproducible evaluation of histopathology classification models released by Dartmouth-Hitchcock Medical Center. The classification task focuses on the clinically important binary distinction between hyperplastic polyps (HPs) and sessile serrated adenomas (SSAs), a challenging problem characterized by substantial inter-pathologist variability, where HPs are typically benign and SSAs represent precancerous lesions requiring closer clinical follow-up. Histologically, HPs exhibit superficial serrated architecture and elongated crypts, whereas SSAs are characterized by broad-based, often complex crypt structures with pronounced serration. A conventional ResNet-18 architecture and its spiking counterpart are evaluated under matched training and inference to isolate the effect of spiking computation. Model’s performance is quantified using the area under the receiver operating characteristic curve (ROC-AUC), yielding 0.817 for the conventional CNN and 0.812 for the SNN. This comparison enables a direct assessment of how spiking computation influences discriminative performance in HPs versus SSAs binary classification and provides a benchmark reference for SNNs on the MHIST dataset. The code is publicly available at https://github.com/qug125/snn-crcp.

## I. Introduction

Digital pathology has become an integral component of modern pathology workflows, enabling large-scale computational analysis of histopathological images. In this domain, convolutional neural networks (CNNs) are widely used for tumor classification tasks, particularly in tile-based image analysis [1]. In colorectal pathology, standardized benchmarks such as A Minimalist Histopathology Image Analysis Dataset (MHIST) released by the department of pathology and laboratory medicine at Dartmouth-Hitchcock Medical Center have further supported reproducible evaluation for histopathology classification [2].

As pathology pipelines scale, there is increasing interest in alternative computing paradigms that may offer different performance-efficiency tradeoffs compared to conventional artificial neural networks. Spiking neural networks (SNNs) represent one such paradigm, introducing event-driven computation through discrete spike-based processing. Recent advances in training techniques have made it feasible to train deep SNNs using gradient-based optimization, motivating their consideration for a broader range of imaging tasks [3], [4].

Despite growing interest in spiking computation, its behavior in digital pathology remains insufficiently characterized under standardized benchmark conditions. In particular, while MHIST has been widely adopted for evaluating conventional CNN architectures, SNNs have not been evaluated on this benchmark. To the best of our knowledge, no prior work has conducted a controlled comparison between spiking and non-spiking CNN architectures on MHIST under identical training and evaluation protocols.

In this work, we aim to address this gap by presenting a benchmark-based evaluation of SNNs for binary colorectal serrated polyp subtype classification on MHIST. We performed a controlled comparison between a standard ResNet-18 and its spiking counterpart under matched experimental conditions, isolating the effect of spiking computation. Our results provide empirical insight into the behavior of spiking CNNs in a standardized digital pathology setting and establish a reproducible reference point for future research.

## II. Related Works

This section reviews prior works on CNN-based approaches for binary classification of colorectal serrated polyp subtypes, specifically distinguishing hyperplastic polyps (HPs) from sessile serrated adenomas (SSAs), summarizes benchmarks on the MHIST dataset, and examines the use of SNNs in medical imaging. Together, these studies provide context for evaluating spiking and non-spiking neural network architectures under controlled experimental settings in digital pathology.

### A. Binary Classification of Colorectal Serrated Polyp Subtypes: HPs versus SSAs

Accurate discrimination between HPs and SSAs is a central task in colorectal pathology. These two colorectal serrated polyp subtypes are associated with fundamentally different risks of malignant transformation and therefore require tailored disease surveillance and management strategies [5]. Although both belong to the serrated class of colorectal polyps and share a saw-tooth crypt morphology [5], SSAs are recognized as precursors of colorectal cancer (CRC), whereas HPs are generally regarded as benign lesions [6]. The World Health Organization therefore defines these entities using distinct architectural criteria, with SSAs characterized by distorted, dilated, and abnormally proliferating crypts extending to the crypt base, and HPs showing superficial serration with preserved crypt spacing and basal proliferative zones [5]. Despite these formal definitions, multiple clinicopathological studies have demonstrated that HP versus SSA differentiation remains challenging in routine practice, with substantial interobserver variability and frequent re-classification when additional serial sections are examined, highlighting the intrinsic ambiguity of this diagnostic boundary [5].

Motivated by this diagnostic difficulty and its clinical consequences, deep learning (DL) has been explored as an approach to improve the reproducibility and scalability of colorectal polyp classification. Wei and colleagues evaluated the ResNet for automated classification of four major colorectal polyp types, including tubular adenoma (TA), tubulovillous or villous adenoma (TVA), HP, and SSA, using digital pathology images [7]. Using majority vote from five gastrointestinal pathologists as the ground truth, their model achieved comparable performance to local practicing pathologists on the independent testing sets, demonstrating that CNNs could learn histomorphological patterns sufficient to discriminate between HPs and SSAs at a clinically relevant level of performance [7]. This work provided one of the first large-scale validations of CNN-based systems for colorectal polyp subtype classification [7].

Subsequent studies using alternative digital pathology pipelines have further supported the feasibility of automated HP versus SSA classification. Byeon et al. trained DeseNet-161 and EfficientNet-B7 on digital photographs of colonoscopy specimens encompassing four diagnostic categories, including TA, traditional serrated adenoma (TSA), HP, and SSA, and reported per-class classification for both HP and SSA, with area under the receiver operating characteristic curve (AUC-ROC) exceeding 0.99 [8]. Collectively, these findings establish HP versus SSA classification as a clinically important and computationally tractable benchmark for evaluating different neural network architectures in digital colorectal histology.

### B. MHIST Dataset and CNN Benchmarks

Wei et al. introduced the MHIST benchmark to support controlled and reproducible evaluation of DL models for binary colorectal serrated polyps subtype classification, focusing on discrimination between HPs and SSAs, thereby supporting systematic methodological experimentation in digital pathology. Rather than emphasizing whole slide imaging (WSI) pipelines, MHIST was designed as a simplified testbed that enables systematic comparison of model architectures using standardized inputs and training protocols [2].

Using MHIST, they demonstrated that CNNs provide strong baseline performance for binary classification of colorectal serrated polyp subtypes (*i*.*e*., HPs versus SSAs) and identified ResNet-18 as a particularly effective reference architecture. Their analysis showed that deep CNN variants do not consistently improve performance on MHIST and may be prone to overfitting, highlighting the suitability of ResNet-18 for controlled architectural comparisons. As a result, subsequent use of MHIST has primarily focused on evaluating conventional CNN-based models under standardized experimental conditions [2].

Despite its role as a benchmark for CNN evaluation, prior work on MHIST has been limited to non-SNNs and has largely emphasized classification accuracy under standard inference settings. The behavior of alternative neural paradigms, such as SNNs, has not been systematically examined on MHIST. This gap motivates the use of MHIST as a testbed for controlled comparisons between spiking and non-spiking neural network architectures in binary colorectal serrated polyp subtype classification of HPs and SSAs.

### C. SNNs in Digital Pathology

SNNs are a class of biologically inspired models that represent and process information using discrete spike events over time, offering an alternative to conventional artificial neural networks [3], [4]. Recent advances in training techniques, including surrogate gradient methods, have enabled SNNs to be applied to a growing range of vision and medical imaging tasks [9].

Within digital pathology, a small but emerging body of work has explored the feasibility of SNN-based approaches for histopathological image analysis. Prior studies have applied spiking models to cancer classification tasks across multiple tissue types, including breast cancer, oral squamous cell carcinoma, skin cancer, CRC, and osteosarcoma [10]–[13].

In parallel, spiking-inspired mechanisms have also been incorporated into standard CNNs, for example through spiking attention modules, to improve interpretability and feature localization, rather than to replace non-spiking computation [14]. While such methods demonstrate the potential utility of spiking dynamics within pathology pipelines, they do not constitute end-to-end spiking CNNs and are evaluated under heterogeneous experimental settings.

Despite these advances, existing SNN-based pathology studies are largely conducted on custom datasets with task-specific preprocessing, architectures, and evaluation protocols. To date, SNNs have not been systematically evaluated on binary colorectal serrated polyp subtype classification (*i*.*e*., HPs versus SSAs) benchmarks such as MHIST, nor compared directly against non-spiking CNN counterparts under identical training and inference conditions. To the best of our knowledge, SNNs have not been evaluated on the MHIST benchmark. This gap motivates a controlled comparison between spiking and non-spiking CNN architectures on MHIST to assess the practical behavior of spiking computation in binary classification of colorectal serrated polyp subtypes, specifically distinguishing HPs from SSAs.

## III. Materials and Methods

This study evaluates spiking and non-spiking convolutional neural networks under a controlled experimental framework for binary colorectal serrated polyp subtype classification using the MHIST dataset, a standardized benchmark for reproducible histopathology image analysis. Figure 2 illustrates the experimental pipeline, highlighting shared data preprocessing, training, model selection, and evaluation procedures, with differences restricted to the neural computation paradigm.

**Fig. 1.**
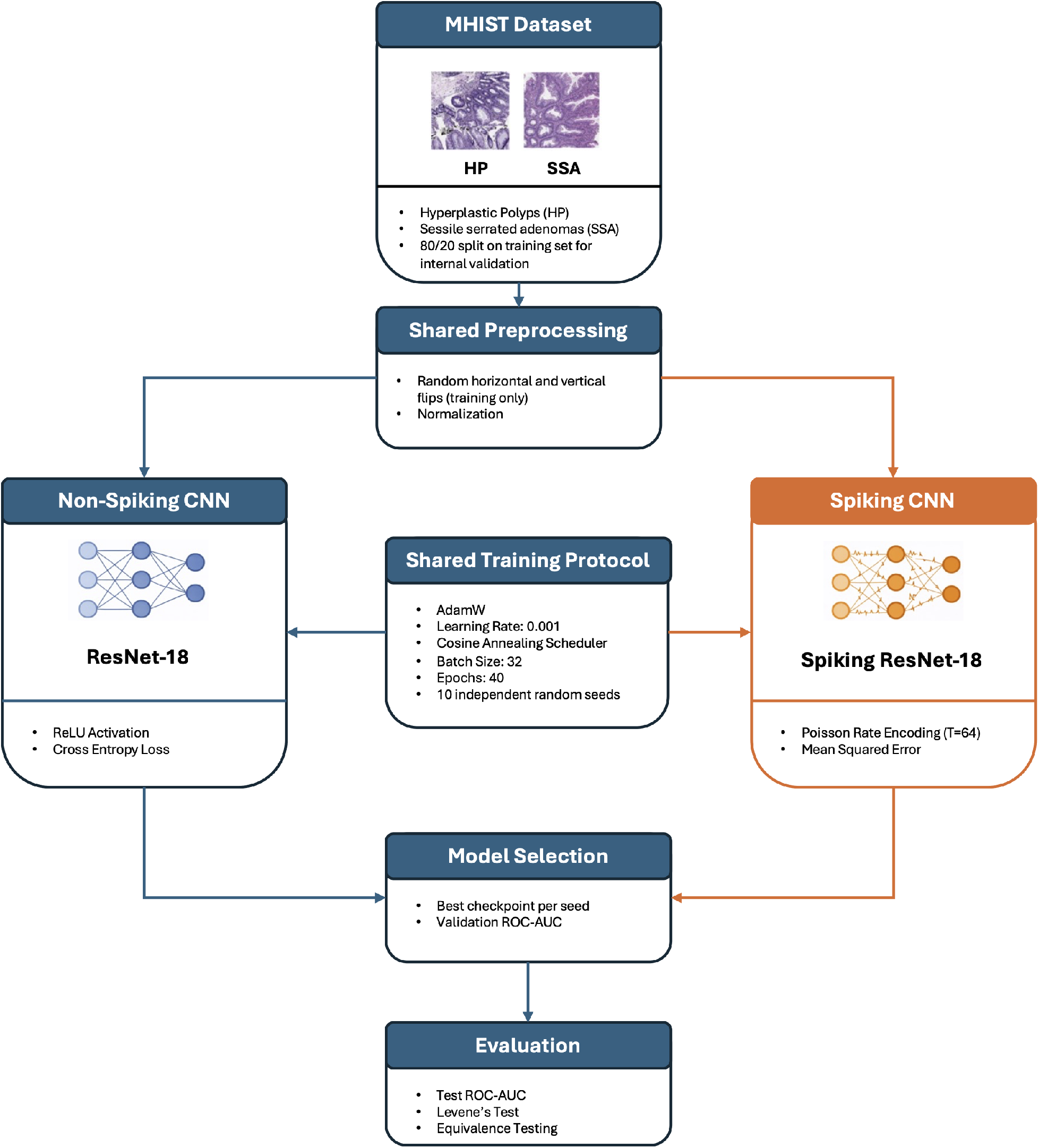
Overview of the experimental pipeline used for controlled comparison of non-spiking and spiking convolutional neural networks on the MHIST dataset. Both models share identical data splits, preprocessing, training protocol, model selection, and evaluation procedures. The two branches differ only in activation mechanisms and temporal spike-based computation.

**Fig. 2.**
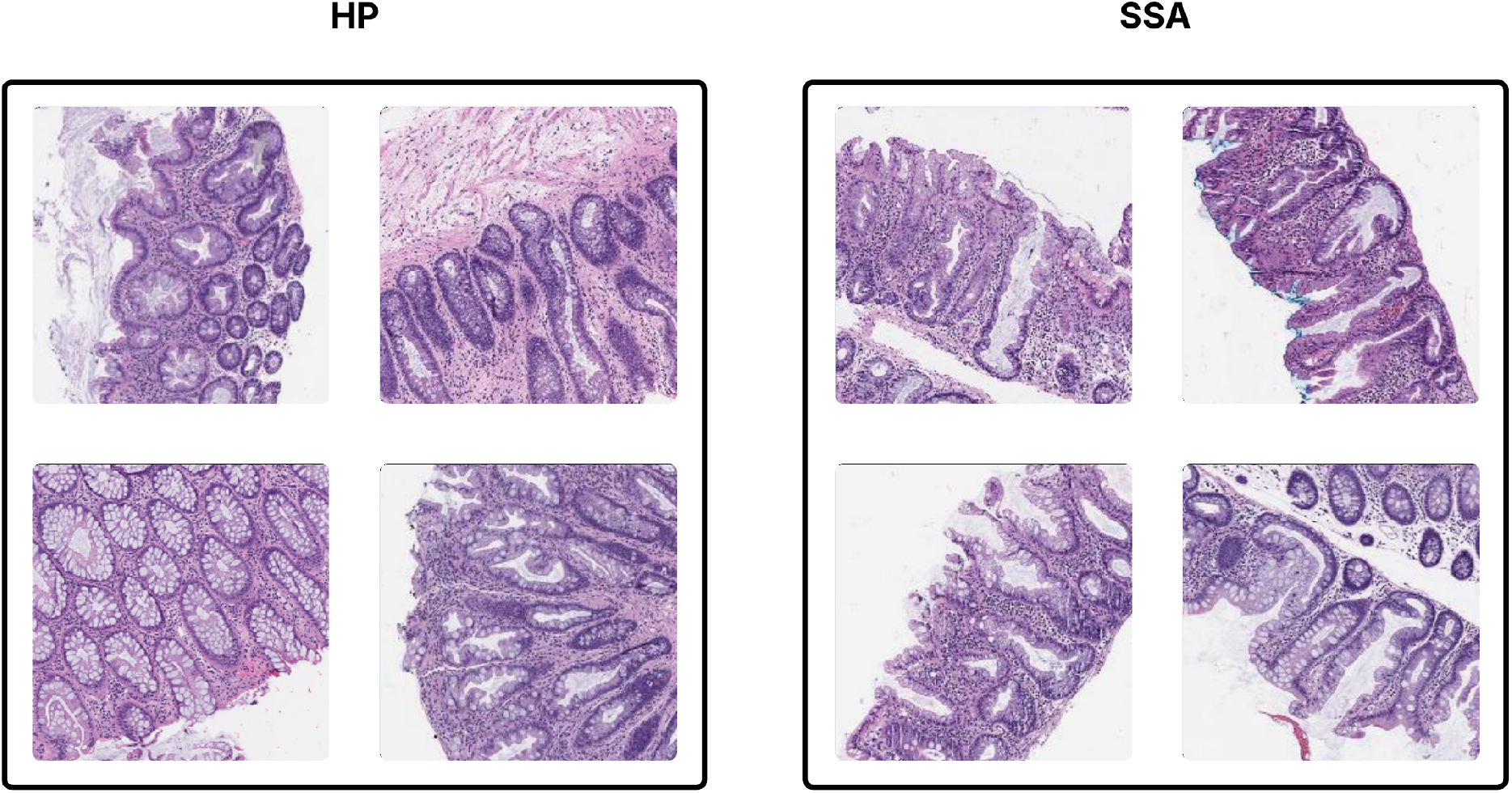
Representative histopathology image tiles from the MHIST dataset. Example tiles from the HP and SSA classes are shown.

### A. Dataset

We evaluate spiking and non-spiking CNNs using the MHIST dataset, a standardized benchmark for binary colorectal serrated polyp subtype classification. MHIST consists of H&E-stained histopathology image tiles labeled as either HP or SSA, enabling a clinically relevant binary classification task. Representative examples of each of these classes are shown in Figure 2.

Although the original MHIST benchmark does not define a dedicated validation split, we construct an internal validation set by partitioning the original training data using an 80/20 split. This validation set is used exclusively for model selection and is held fixed across all experiments. All models are trained on the remaining portion of the original MHIST training set and evaluated on the original MHIST test set. Identical training, validation, and test partitions are applied to both spiking and non-spiking models to ensure a controlled and fair comparison under the dataset’s inherent class imbalance. The resulting class distribution across splits is summarized in TABLE I.

**TABLE I:** Distribution of image tiles in the MHIST dataset across training, validation, and test splits for the two diagnostic classes (HP and SSA). The validation set is derived from the training data and preserves the original class imbalance.

|  | Train | Validation | Test | Total |
| --- | --- | --- | --- | --- |
| HP | 1227 | 318 | 617 | 2162 |
| SSA | 513 | 117 | 360 | 990 |
| **Total** | 1740 | 435 | 977 | 3152 |

### B. Data Preprocessing

All histopathology image tiles were processed using a consistent preprocessing within each model family. All MHIST tiles are provided at a native resolution of 224 *×* 224 pixels and were used directly without resizing.

For non-spiking ResNet-18 models, input images were normalized using standard ImageNet channel-wise mean and standard deviation normalization, consistent with common practice for ResNet-based architectures. For spiking ResNet-18 models, pixel intensities were scaled to the [0, 1] range. This scaling is required for rate-based Poisson spike encoding [15], where pixel intensities paramaterize spike generation probabilities. Aside from this normalization difference, no additional preprocessing differences were introduced.

The original MHIST paper does not explicitly specify the data augmentation procedures used. In our study, data augmentation was therefore limited to simple stochastic geometric transformations consisting of random horizontal and vertical flips applied during training. These augmentations were applied identically across all models.

### C. Model Architecture

We evaluate both a conventional ResNet-18 and its spiking counterpart to enable a controlled comparison between non-spiking and spiking computation in the colorectal serrated polyp subtype classification. Across all experiments, architectural differences were restricted to the neuron activation mechanisms, while the overall network topology was preserved.

1. *ResNet-18 Baseline:* As a non-spiking baseline, we employ a standard ResNet-18 architecture for image classification. ResNet-18 has been widely adopted in digital pathology and serves as a strong baseline on the MHIST benchmark. The network processes static image inputs using continuous-valued activations and is trained using conventional backpropagation. No architectural modifications were made to the model.
2. *Spiking ResNet-18:* For the spiking model, we use the built-in Spiking ResNet-18 implementation provided by the SpikingJelly framework. This implementation follows the standard ResNet-18 architecture while replacing conventional activation functions with spiking neuron units and introducing temporal dynamics through discrete simulation timesteps. Network depth, convolutional filter sizes, and residual connections are identical to the non-spiking ResNet-18 baseline.

Spiking neurons were modeled using integrate-and-fire (IF) neurons. Training was performed using the arctangent surrogate function. Static input images were converted into spike trains using rate-based Poisson encoding and processed over *T* discrete timesteps, yielding a continuous-valued representation used for loss computation and evaluation.

### D. Training Protocol

All models were trained using a unified training protocol to ensure comparison between non-spiking and spiking models. Non-spiking and spiking models shared identical optimization settings, differing only in the presence of temporal simulation of spiking neurons. All training configurations are shown in TABLE II.

**TABLE II:** Model architecture and training configuration used for spiking and non-spiking experiments on the MHIST dataset.

| Configuration | ResNet18 | SNN-ResNet18 |
| --- | --- | --- |
| Input Resolution | $224 \times 224$ | $224 \times 224$ |
| Input Normalization | ImageNet mean/std | [0, 1] scaling |
| Temporal Encoding | None | Poisson (rate-based) |
| Number of Timesteps ($T$) | – | 64 |
| Neuron Model | ReLU | IF neuron |
| Surrogate Gradient | – | Arctangent |
| Temporal Aggregation | – | Mean spike rate |
| Loss Function | Cross-Entropy | Mean Squared Error |
| Optimizer | AdamW | AdamW |
| Initial Learning Rate | $1 \times 10^{-3}$ | $1 \times 10^{-3}$ |
| Learning Rate Schedule | Cosine Annealing | Cosine Annealing |
| Batch Size | 32 | 32 |
| Training Epochs | 40 | 40 |
| Number of Seeds | 10 | 10 |

Models were optimized using the AdamW optimizer with an initial learning rate of 1 *×* 10^−3^. A cosine annealing learning rate schedule was applied with updates once per epoch, providing a smooth decay appropriate for the limited number of training epochs and the computational cost of spiking models. Model selection was performed based on validation set performance, with the checkpoint achieving the best validation ROC-AUC selected for evaluation.

To account for stochasticity in weight initialization, data shuffling, and spike generation, all experiments were repeated across ten independent random seeds. Each seed defines a complete training run, including weight initialization and mini-batch ordering. Model selection was performed independently for each seed using the fixed validation set, and final performance is reported as the mean and standard deviation of test ROC-AUC across seeds.

### E. Model Evaluation

Model performance was evaluated on the held-out MHIST test set using ROC-AUC, consistent with the original MHIST benchmark and appropriate for the class-imbalanced setting. The test set was not used during training or model selection.

To assess whether performance differences between spiking and non-spiking models were statistically meaningful, equivalence testing was performed on per-seed AUC values using the two one-sided tests (TOST) procedure.

## IV. Results

We evaluate the performance of spiking and non-spiking ResNet-18 models on the MHIST dataset under identical training and evaluation conditions. All results are reported on the held-out MHIST test set using ROC-AUC, averaged across ten independent random seeds. Mean performance and variability are reported as mean *±* standard deviation (sd).

### A. Overall Performance Comparison

TABLE III summarizes the test ROC-AUC results for spiking and non-spiking models trained from scratch. Across seeds, both models demonstrate comparable performance on the MHIST test set. The non-spiking ResNet-18 achieves a mean *±* sd ROC-AUC of 0.817 *±* 0.014, while the spiking ResNet-18 achieves a mean *±* sd ROC-AUC of 0.812 *±* 0.023.

**TABLE III:** Test ROC-AUC performance (mean *±* sd) across ten random seeds for spiking and non-spiking models trained from scratch on the MHIST dataset.

| Model | ROC-AUC (mean ± sd) |
| --- | --- |
| ResNet-18 | $0.817 \pm 0.014$ |
| SNN-ResNet-18 | $0.812 \pm 0.023$ |

Although the non-spiking model achieves a slightly higher mean ROC-AUC, the spiking model exhibits overlapping performance distributions across seeds. This suggests that spiking computation can achieve competitive performance on colorectal serrated polyp subtype classification despite the use of spike-based temporal encoding and surrogate gradient training.

### B. Variability Across Random Seeds

To assess the stability of spiking and non-spiking models under stochastic training conditions, we evaluated variability in test ROC-AUC across the ten independent random seeds for each architecture. Figure 3 illustrates the distribution of test ROC-AUC values for the SNN-ResNet-18 and ResNet-18 models. While both models achieved comparable median performance, the spiking model exhibited visibly greater dispersion across runs.

**Fig. 3.**
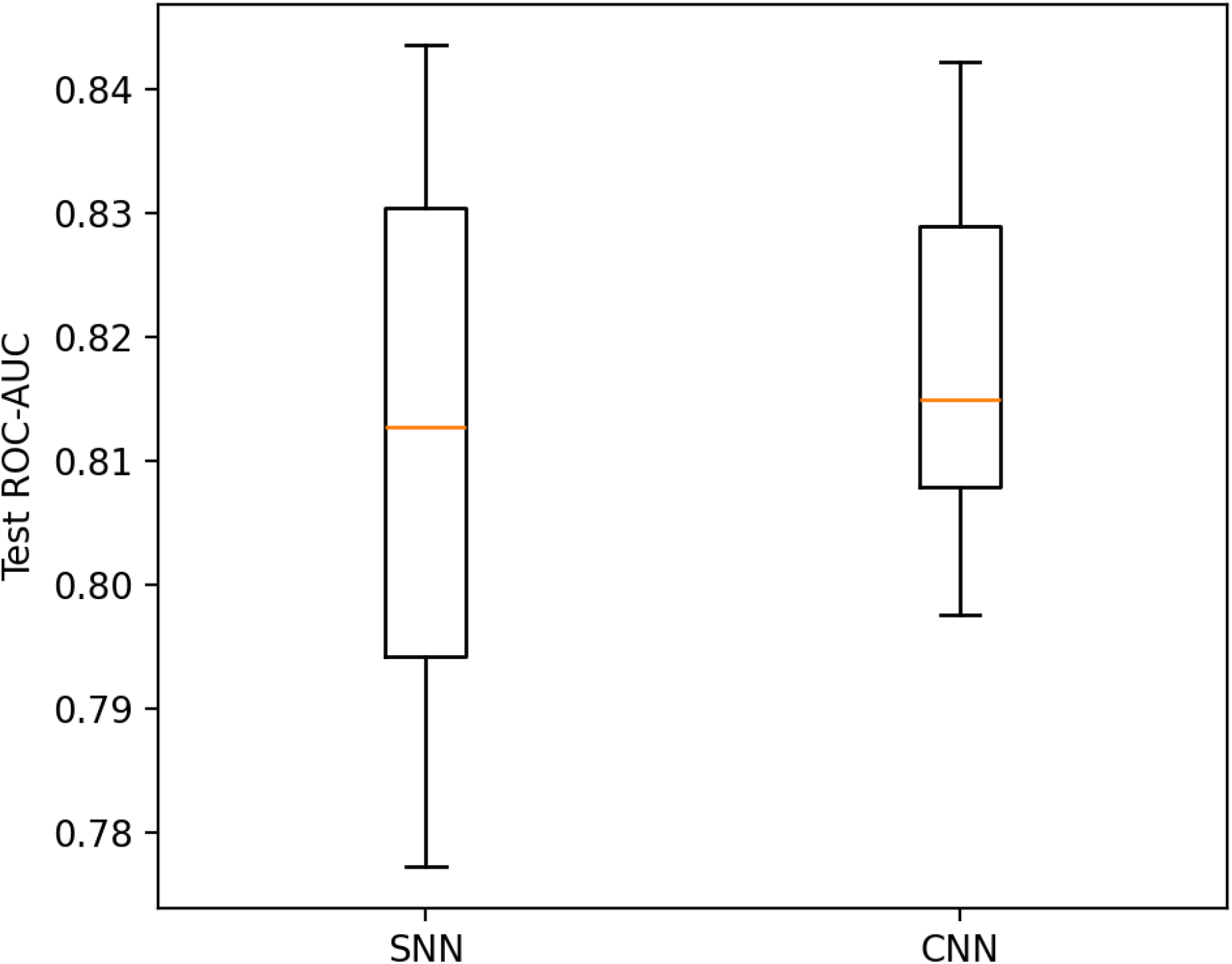
Boxplot of test ROC-AUC across ten independent random seeds for SNN-ResNet-18 and ResNet-18 trained from scratch on the MHIST dataset. While median performance is comparable between models, the spiking model demonstrates increased dispersion across runs, reflecting higher sensitivity to stochastic training effects.

To formally quantify differences in variability, we applied Levene’s test for equality of variances using a median-centered formulation, which is robust to non-normality and appropriate for performance distributions in machine learning experiments. Levene’s test indicated a statistically significant difference in variance between the two models (*p* = 0.0156) at a significance level of *α* = 0.05, rejecting the null hypothesis of equal variance.

Consistent with this result, the sd of test ROC-AUC across seeds was higher for the spiking model compared to the non-spiking baseline. This increased variability is expected given the additional sources of stochasticity introduced by spike-based Poisson encoding and surrogate gradient training. Importantly, despite this higher variance, the overall performance distribution of the models remained overlapping, and no systematic degradation in median performance was observed for the spiking model.

These findings highlight that while spiking neural networks can achieve competitive classification performance on static histopathology images, their training dynamics exhibit greater sensitivity to random initialization and sampling effects compared to conventional CNNs. This behavior underscores the importance of multi-seed evaluation when benchmarking spiking models and motivates reporting performance as distributions rather than single-point estimates.

### C. Statistical Equivalence Analysis

To determine whether performance differences between spiking and non-spiking models were practically meaningful rather than statistically incidental, we conducted a statistical equivalence analysis using the two one-sided tests (TOST) procedure. Unlike traditional null hypothesis significance testing, which evaluates whether two methods differ, equivalence testing assesses whether observed differences fall within a predefined range of practical irrelevance.

Equivalence testing was performed on paired test ROC-AUC values obtained across ten independent seeds. Pairing was used to control for shared sources of stochastic variability arising from dataset splits and experimental conditions. An equivalence margin of Δ = 0.03 ROC-AUC was selected representing a difference considered negligible relative to the observed run-to-run variability across seeds and consistent with commonly accepted values in medical imaging classification tasks.

For the comparison between SNN-ResNet-18 and ResNet-18, the mean paired difference in test ROC-AUC was − 0.0057. The corresponding 90% confidence interval for the mean difference was [ 0.0214, 0.0101. Since this confidence interval lay entirely within the predefined equivalence bounds of *±* 0.03, both one-sided null hypotheses if the TOST procedure were rejected at *α* = 0.05, and practical equivalence between the two models was established.

These results indicate that, despite differences in computational paradigms and training dynamics, spiking and non-spiking ResNet-18 models achieve comparable discriminative performance on the MHIST dataset when trained under matched experimental conditions.

### D. Ablation Study

To better understand the observed performance differences between spiking and non-spiking networks, we conducted an ablation study examining the impact of spike-based temporal encoding and simulation length while holding network architecture, optimization strategy, and data splits constant. All ablations were performed using models trained from scratch, as pretrained initialization under Poisson encoding was found to introduce additional confounding effects and was therefore excluded from the primary analysis.

#### a) Effects of Spike-Based Temporal Encoding

The primary ablation compares a conventional ResNet-18 operating on static image inputs to its spiking counterpart, which processes the same images through Poisson rate-based encoding over multiple discrete timesteps. Aside from the introduction of spiking neurons and temporal simulation, the two models share identical network topology, optimizer configuration, learning rate schedule and training protocol. The comparison isolates the effect of spike-based computation and temporal dynamics on classification performance.

Across all dataset configurations and random seeds, the spiking model exhibited slightly lower mean ROC-AUC compared to the non-spiking baseline, with increased variability across runs. Statistical equivalence testing demonstrated that the observed performance difference fell within a predefined margin of practical equivalence, indicating that the two models achieve comparable discriminative performance on the task. These findings suggest that, under the evaluated conditions of static histopathology image classification, spike-based temporal encoding neither provides a systematic performance advantage nor results in substantial performance degradation relative to conventional convolutional processing.

#### b) Effects of Temporal Simulation Length

We further examined the influence of the number of simulation timesteps (*T*) used in the spiking network. Preliminary experiments with shorter temporal windows, such as *T* = 4 and *T* = 8, resulted in unstable training dynamics and degraded validation performance, likely due to insufficient spike statistics for reliable rate estimation. Increasing the simulation length to *T* = 32 yield substantially more stable optimization behavior and improved validation ROC-AUC, while further increases to *T* = 64 provided further improvements, however at a computational cost.

#### c) Interpretation

Together, these ablations indicate that the performance gap between spiking and non-spiking models on MHIST is largely attributable to the constraints imposed by spike-based temporal encoding rather than architectural or optimization differences. While spiking networks can achieve performance comparable to conventional CNNs when appropriately trained, the additional time dimension introduces variance and computational overhead without clear performance gains for static image classification tasks.

## V. Conclusion

In this work, we presented the first controlled benchmark evaluation of SNNs on the MHIST dataset for binary colorectal serrated polyp subtype classification (*i*.*e*., HP versus SSA). By comparing conventional ResNet-18 architecture with its spiking counterpart under matched training, optimization, and evaluation protocols, we isolated the impact of spike-based computation on a clinically relevant digital pathology task.

Across ten independent random seeds, spiking and non-spiking models achieved comparable test performance with overlapping ROC-AUC distributions. While the spiking model exhibited higher variability across runs, statistical equivalence testing using the TOST procedure demonstrated that the observed performance differences fell within a predefined margin of practical irrelevance. These results indicate that when trained from scratch under appropriate conditions, SNNs can achieve discriminative performance comparable to conventional CNNs for static histopathology image classification.

Our ablation analysis further suggests that primary factors influencing spiking model behavior are related to spike-based temporal encoding and simulation length rather than architectural capacity. While the increasing number of timesteps improves stability and performance, it also introduces additional computational overhead without clear performance gains over conventional convolutional processing for static image inputs. This highlights an important trade-off between biological plausibility, computational cost, and practical utility in pathology workflows.

Several limitations of this study should be acknowledged. First, we focused on rate-based Poisson encoding for static images, which may not fully exploit the potential advantages of spiking computation. Second, pretrained initialization was excluded from the primary analysis due to confounding effects under spike-based encoding. Finally, evaluation was limited to patch-level classification benchmark and extensions to WSI pipelines and energy-efficiency analyses remain important directions for future work.

Overall, this study established MHIST as a viable benchmark for evaluating SNNs in digital pathology and provides a reproducible reference for future comparisons. Our findings suggest that SNNs can serve as competitive alternatives to conventional CNNs in binary colorectal serrated polyp subtype classification, while also underscoring the importance of multi-seed evaluation and carefully statistical analysis when bench-marking spike-based models. Future work will explore alternative encoding strategies, event-based inputs, and hardware-aware evaluations to better characterize the potential advantages of spiking computation in clinical pathology settings.

## Data Availability

All data produced in the present study are available upon reasonable request to the authors

## Acknowledgment

The authors would like to acknowledge the collaborative contributions that made this study possible. QG and NL jointly designed the experimental framework. NL implemented the models and conducted the computational experiments. RX and RB provided clinical and biological insights. NL, QG, RX, and RB jointly wrote and edited the manuscript. All authors contributed equally to this work.

## Notes

### Competing Interest Statement

The authors have declared no competing interest.

### Funding Statement

This study did not receive any funding

### Author Declarations

The study used (or will use) ONLY openly available human data that were originally located at: https://bmirds.github.io/MHIST/ The dataset is de-identified and released with permission from Dartmouth-Hitchcock Health (D-HH) Institutional Review Board (IRB).

